# Exploring the possibility of collaboration for biomedical professionals and traditional healers: A Systematic Review

**DOI:** 10.1101/2023.11.21.23298620

**Authors:** Onaedo Ilozumba, Suyasha Koirala, Anthony Meka, Edmund Ossai, Sopna Mannan Choudhury, Ryan Wagner, Richard Lilford

**Affiliations:** Institute of Applied Health Research, College of Medical and Dental Sciences, University of Birmingham, Edgbaston, Birmingham, B15 2TT, UK; Dhulikhel Hospital, Kathmandu University Hospital (DH-KUH), Dhulikhel, Kavrepalanchok 45200; Programs Department, RedAid Nigeria, Enugu 400102, Nigeria; College of Health Sciences, Ebonyi State University, Abakaliki; MRC/Wits Agincourt Research Unit, University of the Witwatersrand, Johannesburg

## Abstract

**Introduction:** Traditional healers play a crucial role in healthcare provision, particularly in low- and middle-income countries. Thus, there is a global interest in understanding the possibilities for collaboration between traditional healers and biomedical professionals. We believe there is the need for a comprehensive review on collaboration between traditional healers and biomedical professionals. Therefore, the aim of this review is to synthesise the literature on collaboration between traditional healer’s and biomedical professional including relevant interventions.

**Methods:** A systematic review was conducted, utilizing a search strategy in PubMed, Web of Science, SCOPUS, and Google Scholar. Articles addressing collaboration between traditional healers and biomedical professionals were included, with a focus on attitudes, perceptions, interventions, and collaborative models. Data extraction followed a predefined template and the D’Amour et al. framework was employed for analysis.

**Results:** The review identified 29 relevant articles, predominantly conducted in Africa. The majority of studies (n=22) explored attitudes and perceptions, revealing a willingness among traditional healers to collaborate, while biomedical professionals exhibited mixed feelings. Seven studies focused on interventions aimed at fostering collaboration primarily focused on improving referral systems and educational initiatives. These studies found positive outcomes. Examining collaboration through the lens of D’Amour et al.’s framework revealed that trust was a significant barrier to collaboration.

**Conclusion:** This review highlights a willingness to collaborate amongst of traditional healers and biomedical professionals and provides some successful examples of working across systems. It also reveals areas for attention in developing collaborative models of working.

## INTRODUCTION

The World Health Organisation (WHO) provides one of the most widely accepted definitions of a health system. A health system is understood “as comprising all the organizations, institutions and resources that are devoted to producing health actions^1^.” Although this definition is seemingly broad, it excludes traditional medicine and its practitioners. Traditional medicine encompasses a range of medication or procedure-based health care therapies including herbal medicine, naturopathy, ayurveda and traditional Chinese medicine. According to the WHO, 170 of 179 of their member states that responded to a survey, acknowledged the use of traditional and complementary medicine in their countries but only 40 acknowledge them within the health system^2^.

One challenge in recognising traditional medicine is the diversity of providers; traditional healers is an umbrella term which refers to healers with varying types of training and expertise^3^. They are a source of care provision in many countries. In this paper we focus on their use in low- and middle-income countries (LMIC). The popularity of traditional medicine and healers is well studied, and key factors determining their us can be classified as personal, interpersonal, and structural^4–7^. Perhaps due to their long history in communities, traditional healers are often trusted by their clients, even when faced with poor outcomes. There is also a strong community factor in the utilisation of traditional healers with family and friends advocating for the use of traditional healers. For example, children are often taken to traditional healers by their parents^8^.However, in many countries, particularly in LMICs, the use of traditional healers is facilitated by structural problems such as long distances to hospitals, long-waiting times at hospitals, lack of personalised care at health centres and high costs of biomedical treatments^9^.

Whatever the strengths and limitations of traditional healers, there is no evidence that they will decline or vanish – on the contrary, traditional healing is a prominent service even in high-income countries. A question that then arises regards the interface between traditional healers and biomedical services. Could traditional healers be a source of referrals to biomedical professionals? Could biomedical professionals be a source of referrals to traditional healers?

Even though traditional healers and biomedical practitioners usually operate in the same space, their practices are often independent of each other. While collaboration between both sectors is limited, there is evidence that some programs and countries have attempted to improve collaboration between biomedical and traditional medicine practitioners. Recent reviews have examined opportunities for integration of traditional healers in the provision of mental health ^10,11^, HIV/STDs^12,13^ and primary care^14^. However, reviews tend to be focused on specific conditions and we believe there is the need for a comprehensive review on collaboration between traditional healers and biomedical professionals. Therefore, the aim of this review is to synthesise the literature on collaboration between traditional healer’s and biomedical professional including relevant interventions.

We recognise that collaboration comes in different shapers and that there are numerous models which could be used to identify different types of collaboration. In our review we base our concept of collaboration on the framework of D’Amour et al. (2005)^15^. They identified four common types according to degree of integration; sharing, partnership, interdependence, and power. Figure 1 describes these types in more detail.

**Figure 1:**
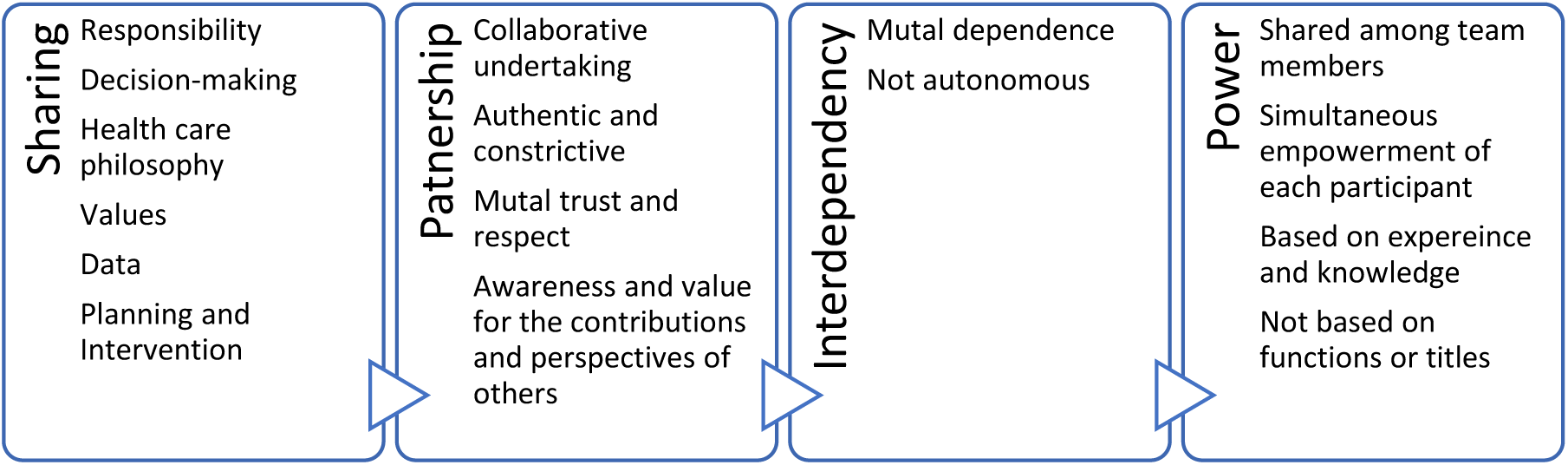
Concepts Related to Collaboration

## METHODS

### Definitions and Theoretical Framework

In this systematic review, we sought to explore the existing evidence on collaboration between traditional healers and biomedical professionals. We will conceptualize traditional medicine according to the WHO definition of traditional medicine as “*the sum total of the knowledge, skill, and practices based on the theories, beliefs, and experiences indigenous to different cultures, whether explicable or not, used in the maintenance of health as well as in the prevention, diagnosis, improvement or treatment of physical and mental illness.”*^2^ While we will work with this definition to select articles got inclusion , we will explore how authors and different cultures define traditional healers.

### Search Strategy

We developed a search strategy (Table 1) which was used in PubMed, Web of Science, Google Scholar, and SCOPUS on 29th March 2022. On 17^th^ May 2023, we conducted an updated search in PubMed, Web of Science and SCOPUS. We did not conduct an updated search on Google Scholar because the system does not allow for refining by specific dates.

**Table 1:** Selected Definitions of Traditional Healers.

| Definitions of Traditional Healers | Articles |
| --- | --- |
| Traditional healers who “know the folklore of disease causation, treatment and prevention” and are known by the term ‘bomoh’. Bomoh use a variety of handed-down, traditional methods in diagnosing and treating patients including “herbal remedies, ceremonial rites, incantation, exorcism and sorcery.” | Merriam, S. B., & Muhamad, M. (2012) |
| • Traditional practitioners (ixhwele), who are predominantly men who specialise in the use of herbal medicines<br>• Diviners (Igqira), who are predominantly women who are called by the ancestors to become a diviner to enable them to acquire the knowledge and skills of traditional healing<br>• Faith healers, who are people who use the power of suggestion, prayer, and faith in God to promote healing<br>• Traditional birth attendants, or traditional midwives, who are middle-aged or elderly ladies with no formal training who attend to women during pregnancy, labour, and the post-natal period by using herbs to facilitate delivery or cause bleeding of the uterus post-natally | van Rooyen et al. (2015). |
| Traditional healers are broadly grouped into three types:<br>1. The traditional doctor or inyanga who is typically male and uses herbal and other medicinal preparations for treating disease (or herbalist);<br>2. The isangoma (Zulu) or diviner, usually a woman who operates within a traditional religious supernatural context and acts as a medium with the ancestral shades; and<br>3. The faith healer who integrates Christian ritual and traditional practices. | Mngqundaniso, N., & Peltzer, K. (2008); Keikelame (2015) |
| Kleva: Bislama word for a traditional healer or traditional medical practitioner. The kleva's role is as a benevolent healer, with no connotations of sorcery.<br>Kastom: The word that people in Vanuatu use to characterise their own knowledge and practice in distinction to everything they identify as having come from outside their place.<br>Dokta: Bislama translation of the word "doctor". Kastom dokta refers to a traditional healer in Vanuatu.<br>Traditional healers use either acquired or inherited knowledge, or divinely given the insight to diagnose and treat social and health problems, and they use prayer, massage and leaf medicine for cure and prophylaxis [28]. In Vanuatu, they often combine "botanical expertise with an ability to diagnose illnesses using supernatural powers". They understand that the causative agent is a pathogen, a social taboo or transgression, and the solution is often based on the traditional healer's ethno-botanical and spiritual knowledge. | Viney et al. (2014). |
| Traditional Healer's definition is based on the definition given by the Ethiopian Health system. One type of the THs is Faith Healers, (called Sheekota, in the local language), who is believed to have some spiritual endowment and they use prayers to heal those they believe are suffering from a 'bad spirit' (Personal communication with HEWs). The most known THs are herbalists, followed by bone settlers, and birth attendants, locally called Qorich Aaada, Dhidhibaa and deesisttu, respectively (Personal Observation).<br>Herbalists (called Qoricha Aaada locally) are most common and they use wild plants (roots and leaves of plants) to treat common health problems like intestinal parasites and snake bites etc. Bone settlers (called Dhidhibaa/Dhidibtuu locally) practice realign- ing fractured bones, tooth extraction and cutting out the uvula. There are also traditional birth attendants (called Deesisttu Aadaa locally) who assist a woman during childbirth. | Sima et al. (2019). |
| Indigenous healing practices broadly operate within a holistic framework that is focused on (a) the interrelationship of nature, (b) health and medicine as gifts, (c) mutual respect in relationships with medicine, (d) imbalances leading to disease, and (d) communal aspects of healing. Moreover, Indigenous approaches to health are often holistic in that they embrace deeply held beliefs about the importance of relationships to the spirit world, to the ancestors and future generations, as well as to the physical and natural world in which we live. | Johnson- Jennings (2018) |

Our developed search string was (“traditional healer” OR “traditional medicine practitioner” OR “bone setter ” OR “faith healer” OR “spiritualist” OR “herbalist” OR “herbalists” OR “medical herbalist”). We did not restrict our search to any particular condition since traditional practitioners do not confirm to the bio-medical nosology. We also omitted any references to collaboration in the search string. Since were concerned that this would exclude papers where a collaboration was couched in different language. We also did not include any language or date restrictions.

### Article Selection

OI, SK, EO and independently screened titles and abstract for full-text reading. OI and SK screened all titles and articles, EO screened 50% of all first stage titles and abstracts. We included articles which addressed perceptions on collaboration between traditional healers and biomedical professionals as well as those that evaluated interventions or programs aimed at facilitation or improving collaboration between traditional healers and biomedical health professionals. Studies were excluded if they focus was restricted to assessing attitudes, practice, or utilisation of traditional healers. We also excluded studies that focused on mental health since, as stated, this topic has been reviewed elsewhere.

We then conducted full text screening of the selected articles. To support our analysis, we developed an extraction template which we piloted with three articles. After discussions and modifications, we used the piloted template to extract data from the included articles. Any differences were decided through discussion and if necessary, we had a fourth reader, SC, available. Data was extracted according to two general study types: i) studies evaluating perceptions and attitudes towards collaboration ii) studies evaluating interventions and programs aimed at fostering, improving, or implementing collaboration processes. We also extracted data on definitions of traditional healers.

## RESULTS

### Included Articles

Our initial search resulted in 4,840 studies, the deletion of duplicates and title and abstract screening 104 potential articles were identified for full screening. After full screening 27 articles were included in the review. We hand searched the references of included articles and found one additional article. Our updated search identified 728 articles published between March 2022 (initial search date) and May 2023 (updated search date) of which 1 was identified for inclusion. (Figure 2: PRISMA flow diagram of update searches.

**Figure 2.**
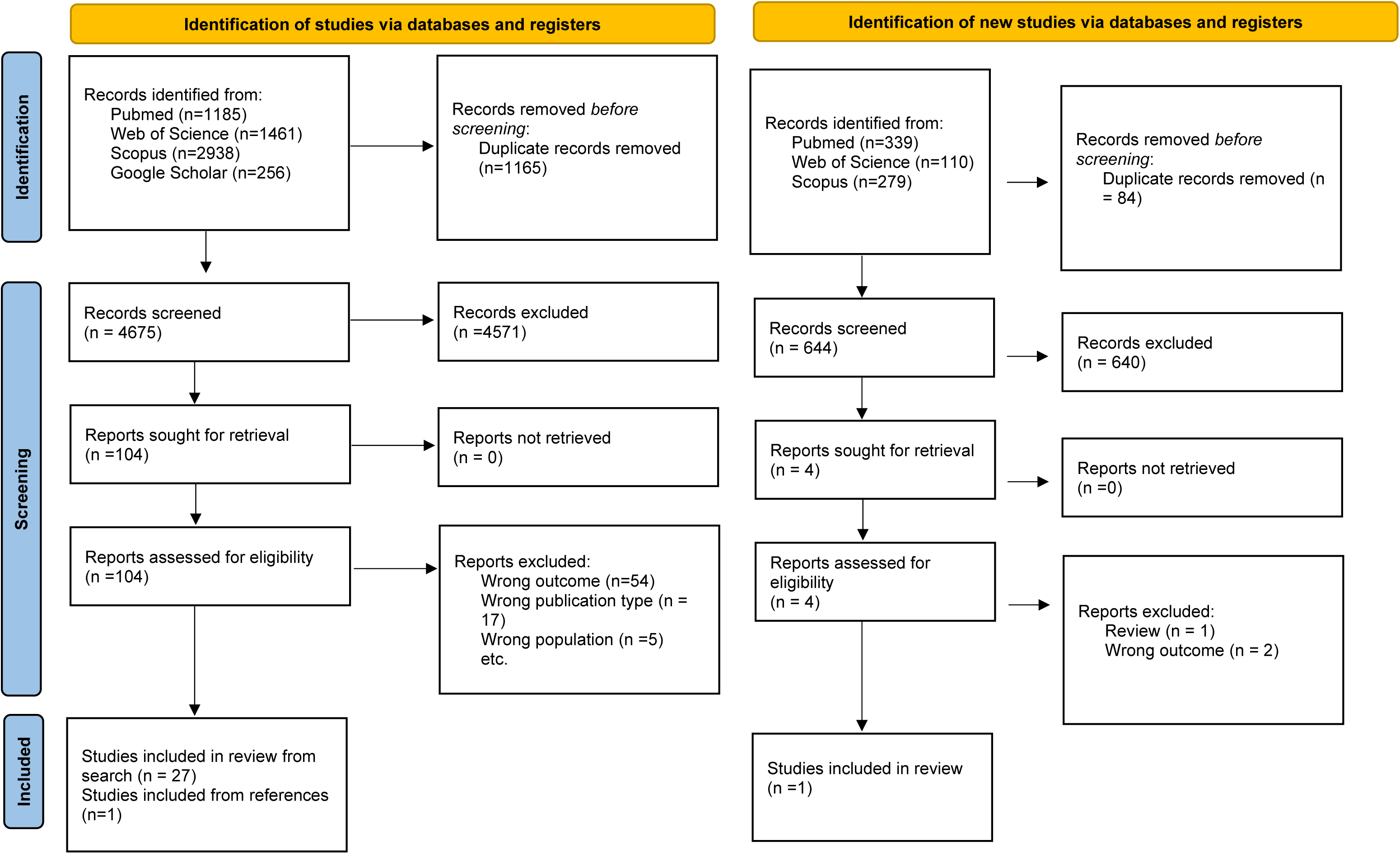
PRISMA Flow Diagram

Of the 29 articles included, 24 were conducted in Africa with the highest number conduced in South Africa (n=8). Thirteen articles focused on the general incorporation of traditional healers into the health care system and 16 articles focused on single condition cases including HIV/AIDS, tuberculosis epilepsy, diabetes, malaria, and cancer.

We present our results in four sections. First, we explore how authors and communities defined traditional healers. Next, we explore articles which discussed attitudes and perceptions towards collaboration. The third section examines articles which described an intervention or approach to collaborating with traditional healers. Finally, we review the presented models of collaboration using the D’Amour et al. (2005) framework.

### Who is a traditional healer?

We sought to understand how our included articles defined traditional healers. We found that included articles took one of three approaches to defining traditional healers. In the majority of the articles (n=14), there was no explicit conceptualization or definition provided for traditional healers^16,17,26–30,18–25^. In eight articles, the authors either used the WHO definition (presented in the methods) or a similar definition of traditional healers as individuals with cultural knowledge of disease and treatment^31–38^. In the final seven articles, traditional healers were described in more culturally specific terms, with attention to distinction between different types traditional healers^39–45^.

In Table 1, we present the specific definitions used in the final group of seven articles

### Perceptions and Attitudes towards Collaboration

The majority of included articles (n= 22) had objectives related to the perceptions and attitude towards collaboration (Table 2). Five studies focused on the opinions and perspectives of traditional healers ^22,39,40,46,47^, six studies exclusively interview or surveyed biomedical professionals ^20,24,36,38,44,48^, eight studies concurrently examined the opinions for biomedical professionals and traditional healers^16,18,21,35,37,41–43^ and two studies included multiple stakeholders^17,33^. Only one study exclusively sought the opinions of policy makers^34^.

**Table 2:** Description of Articles on Perceptions and Attitudes towards Collaboration.

| **Authors (date)** | **Country** | **Study Aim** | **Study Type** | **Data Collection** | **Sample** |
| --- | --- | --- | --- | --- | --- |
| **Primary Care/Health Care system** |  |  |  |  |  |
| **Johnson-Jennings et al. 2018** | USA | *examined whether provider racial concordance (advance practice nurses, physicians, and physician assistants) and ethnic salience of Indigenous patients' effects provider integration of traditional practices with treatment* | Quantitative | *Clinical case vignette and an online survey* | *Study participants consisted of 145 health care providers recruited from 446 IHS service facilities across the United States. 33 American Indian (i.e., racially concordant providers) and 112 non-American Indian (i.e., racially discordant providers). Sixty-five males and 80 females completed the entire survey. Most provider participants were 41 to 55 years of age. Participants included medical doctors (107), nurse practitioners (21), physician assistants (6), and students/others (11).* |
| **Krah et al. 2018** | Ghana | *assessed problems hindering integration and opportunities for integration.* | Qualitative | *In-depth interviews and observations* | *Biomedical professionals' midwives, nurses, and nurse managers and/or hospital coordinators Traditional medicine practitioners and Traditional birth attendants. Patients receiving care either at biomedical health facilities or from traditional healers.* |
| **Kwame Abukari. 2021** | Ghana | *explored perceptions of different actors about integration in Ghana* | Qualitative | *semi-structured interviews* | *60 participants: traditional healers, health care service consumers, and biomedical health care practitioners.* |
| **Lampiao et al. 2019** | Malawi | *evaluated barriers to collaboration between traditional healers and biomedical practitioners.* | Qualitative | *semi-structured interviews* | *In total, 27 semi-structured interviews and 9 focus group discussions (of about 6–8 people) with biomedical professionals (9 interviews, 6 focus groups); NGO sector (1 interview); traditional practitioners (2 interviews; 4 focus groups); and health service users recruited from the sub-district clinics (15 interviews).* |
| **Peltzer K and Khoza LB. 2002** | South Africa | *assessed attitudes and knowledge of traditional healing among nurse practitioners in the Northern Province in South Africa.* | Quantitative | *Questionnaire* | *84 nurse practitioners; 20 were discarded because their questionnaires were not complete. The sample consisted of nurse practitioners working at 9 health centres and 14 clinics of the Lowveld and Northern regions of the Northern Province. The nurses were 78 female (92.9%) and 6 male (7.1%) in the age range of 23 to 58 years (M age 38.0 yr., SD=8.0). All participants were African (black). The number of years of health service ranged from 3 to 27 years, with a mean of 13.0 years (SD=6.9)* |
| **Peu et al. 2001** | South Africa | Identified the attitudes of community health nurses towards integrating traditional healers into primary health care. | Quantitative | Questionnaire | 95 registered nurses who provided care in Odi region of the Northwest Province. |
| **Pinkoane et al. 2008** | South Africa | explored the perceptions and attitudes of policy makers, regarding the incorporation of traditional healers into formal health systems | Qualitative | n/a | Officials in the Provincial Health Departments designated with the job of managing healthcare services in the districts/ regions within the identified provinces; |
| **Suwankhong et al. 2011** | Thailand | examined barriers and facilitator for cooperation between traditional healers and biomedical professionals | Qualitative | participant observations, focus groups, in-depth interviews, and unobtrusive methods. | 60 Participants: traditional healers, customers, community heads, health care providers, health policymakers, and an academic from a local university. |
| **Upvall 1992** | Swaziland | Explored nurses perceptions of collaboration between traditional healers and biomedical professionals. | Qualitative | semi-structured interviews | 65 nurses practising in urban, periurban, and rural areas). Most nurses interviewed represent urban and rural areas almost equally, while periurban areas are under-represented. Also, 85% were employed by government and mission sectors. All nurses interviewed had been practising the profession for at least one year. The length of time practising was related to the nurse's age. The youngest nurse interviewed (age 24) had been practising for 1 year, while the oldest nurse (age 68) had operated her own clinic and practised nursing for 44 years. The other nurse operating her own clinic had been practising for 30 years |
| **Van Rooyen et al. 2015** | South Africa | *explored and describe the collaborative relationship between traditional healers allopathic and traditional health practitioners regarding the legalisation of traditional healing, and these health practitioners' views of their collaborative and professional relationship, as role-players in the healthcare delivery landscape* | Qualitative | *Focus group discussions and unstructured individual interviews* | *28 participants: 10 allopathic health practitioners, 14 traditional healers and 4 traditional healers who are also allopathic health practitioners. Participants were mostly black people (n = 7) and white people (n = 2) with only one mixed race participant, and 9 of the participants were female. The participants' work experience varied from 13 years to 41 years of working in several healthcare facilities, including different wards in district and regional hospitals and psychiatric clinics. Group Two consisted of 14 (n = 14) traditional healers, consisting of 4 diviners, 4 herbalists, 3 traditional surgeons, and 3 birth attendants. The participants were mostly male (n = 10), practicing in local urban, rural and township areas. The participants' level of education varied from illiterate to being in possession of a Doctoral Degree. Lastly, Group Three consisted of 4 (n = 4) participants: 3 registered nurses, and 1 enrolled nurse, who are also traditional healers. All participants were female and black, with work experience as allopathic practitioners ranging from 13 to 31 years and as a traditional practitioner from 3 to 9 years.* |
| **Hoyler et al. 2016** | Guatemala |  | Qualitative |  |  |
| **HIV** |  |  |  |  |  |
| **Burnett et al. 1999** | Zambia | *to examine knowledge and beliefs of traditional healers and formal health workers working in geographical proximity and plan a programme of workshops to encourage their collaboration.* | Qualitative | *Semi-structured interview* | *Thirty-nine traditional healers and 27 formal health workers were interviewed. Of the healers, 22 were male and 17 were female, whilst the formal health workers comprised four men and 23 women. The ages of the healers ranged from 22 to 82 years, with a median of 48. Formal health workers' ages ranged from 25 to 48 years, with a median of 34. Only three traditional healers had attended school beyond Grade 12 (the final year of secondary schooling) and 12 had received no formal education. All the formal health workers had completed further education.* |
| **Madiba (2010)** | Botswana | to determine biomedicine health practitioners' views on collaboration with traditional health practitioners and identify ongoing collaboration activities acceptable approaches for collaboration. | Quantitative | questionnaire | The response rate from biomedicine health practitioners was 100%. Of the BHPs who completed the survey, 36 (60%) were female and 24 (40%) male and their ages ranged from 25 to 50 years. BHPs comprised of 42 (70%) nurses, 5 (5, 8%) medical practitioners, 5 (5, 8%) pharmacists, 7 (12%) were laboratory staff, and only 1 (2%) was a social worker. The demographic profile of BHPs in the study sample is consistent with the distribution of health care practitioners in the country; there are more nurses than any other professional category. |
| **Mendu and Ross (2019)** | South Africa | To obtain information on (1) the views and experiences of biomedical healthcare practitioners and African traditional healers regarding working together as part of the multidisciplinary HIV and AIDS care team; (2) the two groups' knowledge about each other's methods of prevention, diagnosis and treatment of HIV and AIDS; (3) factors perceived by biomedical and traditional practitioners to impede the integration of the two groups from working together; and (4) recommendations for facilitating the integration of the two groups within an HIV and AIDS multidisciplinary team. | Qualitative | Semi-structured interviews | The sample comprised 10 biomedical healthcare practitioners and 10 traditional healers. The sample of biomedical healthcare professionals included 2 medical doctors, 3 nurses, 2 social workers and 3 pharmacists. There were 2 male and 8 female biomedical healthcare practitioners in the sample, with 8 black participants and 2 white participants. Furthermore, the work experience of the sample ranged from 3 to 42 years. The participants were employed by the Department of Health and had direct contact with patients. The sample of African traditional healers incorporated 6 males and 4 females, all of whom were black, with practice experience that ranged between 6 and 36 years. In terms of ethnicity, the sample included five Zulu, two Sotho, one Ndebele, one Shona and one Setswana participant, which enriched the diversity of responses obtained. |
| **Mngqundaniso and Peltzer (2008)** | South Africa | *investigated the role of traditional healers in sexually transmitted infections including HIV and AIDS and collaboration between the traditional and biomedical health care systems as seen by nurses and traditional healers.* | Qualitative | Semi-structured interviews | *15 professional nurses and 15 traditional healers. Mean age 45 (SD 12.8 years, ranged from 22 to 84 years). All nurses were registered professionals from selected primary care clinics in KwaZulu-Natal. Traditional healers included those registered with the healer's council and not registered.* |
| **Viney et al. (2014)** | Vanuatu | *to assess the knowledge, attitude, and practice of traditional healers towards TB in Vanuatu.* | *mixed methods study design (qualitative and quantitative methods).* | semi-structured questionnaire | *Nineteen healers were interviewed.* |
| **Sima et al. (2019_** | Ethiopia | *evaluated the role of THs in the detection and referral of active TB cases in a pastoralist community.* | Qualitative | semi-structured questionnaire | *Twenty-two THs were included in this pilot intervention study. However, only eight of them were available for interview after 2 years. The rest were unavailable due to seasonal migration and instability in the district during the study period.* |
| **Others** |  |  |  |  |  |
| **Merriam et al. (2012)** | Malaysia | to explore how these two systems might be brought closer together in addressing the cancer. | Qualitative | Semi-structured interviews and focus group discussions | 14 traditional healers and 12 medical professionals. Of the 14 participants, six practice in and around Kuala Lumpur, and eight practice in rural areas in the north-western, southern, and north-eastern parts of Peninsular Malaysia. Ten of the 14 THS are men, age ranges from 43 to 80 years, and years in practice range from 11 to 48 years. With regard to type of practice, one is a homeopathic healer, three are known as Islamic healers, one is a herbalist, and nine identify as 'traditional healers' using a variety of spiritual/religious methods, herbs, flowers, fruits and roots. One has had some training in acupuncture, which he occasionally incorporates into his treatment. One woman, Lena, channels a medical doctor, and one man, Aziz, does invisible 'surgery' in his practice. Of the 12 BHPs, five are from the Kuala Lumpur area and seven are from rural areas. Oncologists, radiologists, and surgeons comprise the sample. Two of the participants are women (both surgeons). |
| **Nkosi et al. (2018)** | South Africa | to explore the perceptions of THPs and ROs regarding referral of cancer patients in a cooperative practice in KwaZulu-Natal | Qualitative | Semi-structured interviews | 28 THPs and 4 Ros. In the THPs category, there were 19 males and 9 females. Their age ranged from 20 to 80 years where the majority was at age 40–60 years. Three had no formal education, 12 had primary school education (Grade 1–7), 12 had high school education (Grade 8–12) and one had a diploma. They indicated that they had prior training in the treatment of HIV and AIDS. The majority was traditional herbalists (18), followed by seven diviners, one spiritual healer and two THPs were both a traditional herbalist and a diviner or a traditional herbalist and a spiritual healer. The THPs' work experience as traditional healers ranged from 1 to 60 years with the majority being between 1 and 40 and a minority above 40 years. The ROs were of different races with English as their first language. Of the four ROs, three were males and one was female. Their age ranged from 36 to 46 years and their work experience as an RO ranged from 5 to 15 years. For reasons of confidentiality, their races and positions could not be disclosed as this could lead to them being identified. |
| **Keikelame et al (2015)** | South Africa | To use Kleinman's Explanatory Models (EMs) framework to explore traditional healers' understandings of epilepsy and its management | Qualitative | Semi-structured interviews and focus group discussions | Two were males and four were females. Their mean age was 65 years and 59 years respectively. Two female healers had been trainees ("umkhwetha") for more than 10 years. Of the remaining four, all were sangomas with the two males being sangomas as well as herbalists. All four had been practising for more than 20 years. Only one female trainee ("umkhwetha") did not reside in the study area. All six healers had treated and cared for a patient with epilepsy. Another female trainee ("umkhwetha") had a daughter who had epilepsy. Among the two male healers, one had a nephew who had died from epilepsy and another had personally experienced an epileptic seizure. Four were registered members of THO. Two had never attended school and four had passed grade six. Four were receiving a government pension grant. One was employed as a cleaner and another, who was a trainee, did not disclose her means of income. Out of nine focus group participants (FGP), six were female sangomas and two males were sangomas as well as herbalists. Only one female was a trainee ("umkhwetha"). The mean age was 63 years for men and 61.3 years for females. Three had never attended school and six had passed grade six. Five were registered members of THO. None of the healers in the individual interviews and focus group had prior knowledge of the national Non-governmental Epilepsy Organization and some had received lay training in HIV/AIDS and TB, but not in epilepsy. |

Across all 22 studies, there was a strong perception that traditional healers were willing to collaborate with biomedical professionals. Some studies highlighted existing evidence of this willingness, for example in Lampiao et al. 2019, traditional healers discussed informal referrals to biomedical health facilities^37^. Traditional healers gave examples of incorporating biomedical medicine, for example a hospitals sending x-rays to bonesetters to support their treatment and traditional healers sending patients with open wounds to the hospital for blood and medications ^16^. However, even among those who favoured collaborations, there were sometime concerns about interactions between traditional and biomedical medication.

Biomedical professionals reported more mixed feelings about collaboration, ranging from positive to very negative or supporting a limited approach. In Peltzer et al. (2002), the majority of surveyed nurses felt that integration of faith healers (98%) and traditional healers (93%) would be beneficial. Peu at al. also had a similarly high approval, with 84% supporting integration of traditional healers^32^. However, in the 16 studies which included biomedical professionals, they expressed concerns about collaboration. These were often related to expressed concerns about traditional healers skills, their training and ability as well as lack of guidance for working with traditional healers^21,37,42,49^.

Two studies highlighted a possible variation in traditional healers and biomedical health professionals’ attitudes towards collaboration. Merriam et al. (2012) found that Islamic healers were the most willing to collaborate and traditional healers with minimal formal education were least interested^41^. In Upvall 1992, nurses working at mission urban hospitals or peri-urban clinics opposed collaboration and believed that traditional healers work was detrimental to patients well-being^36^. However nurses at private practice, rural mission clinics, and non-governmental organizations had more positive responses to the idea of collaborating with traditional healers^36^.

Policy makers, in one, study felt that traditional healers could be important resources in the delivery of care. However, there was a need for education, guidelines, and communication^34^.

However, traditional healers and biomedical professionals were often only willing to collaborate under specific conditions. For traditional healers, these ranged from reciprocated collaboration, respect from biomedical professionals, government recognition to remuneration^39,40^ but traditional healers and biomedical professional identified concerns about loss of revenue for traditional healers as a barrier to collaboration^37,39^. One proposed solution was that traditional healers could be paid for referrals.^39^ In contrast, one traditional healer stated that payment for services could be in violation of their medical philosophy. In Thailand, traditional healers suggested two-way referrals as a collaborative model, although none of them currently collaborated with biomedical professionals^33^. Despite their willingness to collaborate, traditional healers also expressed distrust of biomedical professionals. Reasons stated include that they believe that biomedical clinicians would appropriate their knowledge without acknowledgement or credit ^33^. Biomedical professionals on the other hand most often discussed collaboration as traditional healers referring patients to them. Their conditions were generally related to the need for training of traditional healers as well as limiting the services they can render^20,36^. When biomedical professionals were asked if they would ever refer patients to traditional healers 90% responded negatively^24^.

Language difficulties between formal providers and traditional healers were mentioned as a difficulty ^37,49^. In a somewhat similar finding on the need for familiarity, Johnson-Jennings et al. found that racially concordant/Indigenous providers were more likely to refer their patient to herbal practices, F (1, 89) = 5.99, p = .02, and other traditional healing practices, F(1, 89) = 5.03, p = .03^44^. Krah et al. found that having biomedical health professionals who were seen as outsiders with limited understanding of local traditional medicine practices hampered trust between biomedical professionals and health professionals^16^. Van der Watt et al. (2017) found that human resource for health limitations and geographical constraints were additional barriers to collaboration^19^.

### Collaborative Interventions and Programs

Seven of the included articles described an intervention or program developed to initiate or foster collaboration between biomedical professionals and traditional healers (Table 3). Five of these articles focused on initiating or improving referral by traditional healers to health facilities^26–28,50,51^. Interesting to note that these five articles focused on the detection of suspected infectious diseases (tuberculosis^26,27,50,51^ and trypanosomiasis^28^). The sixth study evaluated an intervention aimed at improving knowledge and prevention of diabetes^30^. The final study explored Ghana’s attempts at integrating traditional healers and biomedical professionals in the health system focusing on high level management rather than results at the treatment level^29^. None of the studies used an experimental design.

**Table 3:**
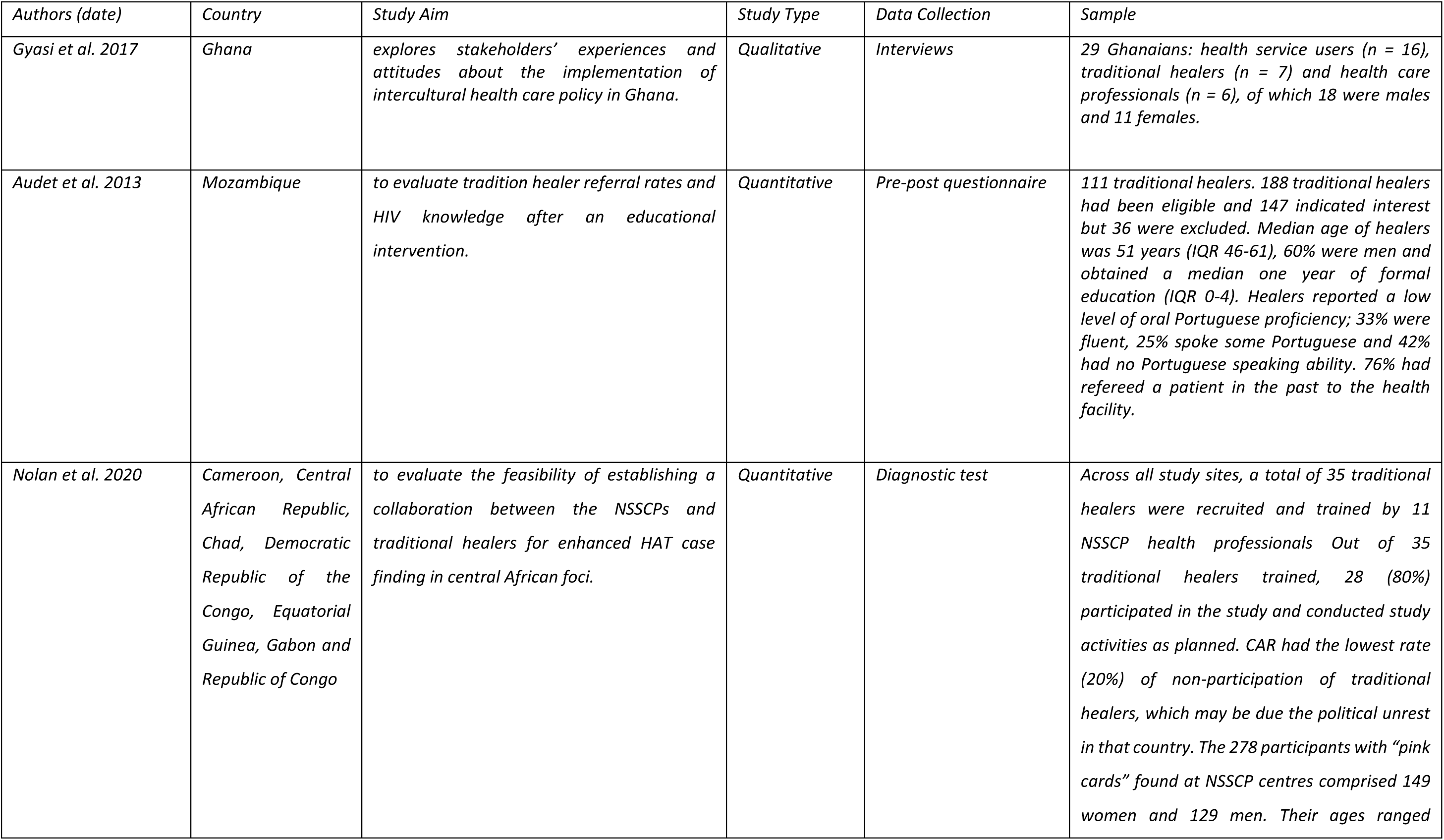
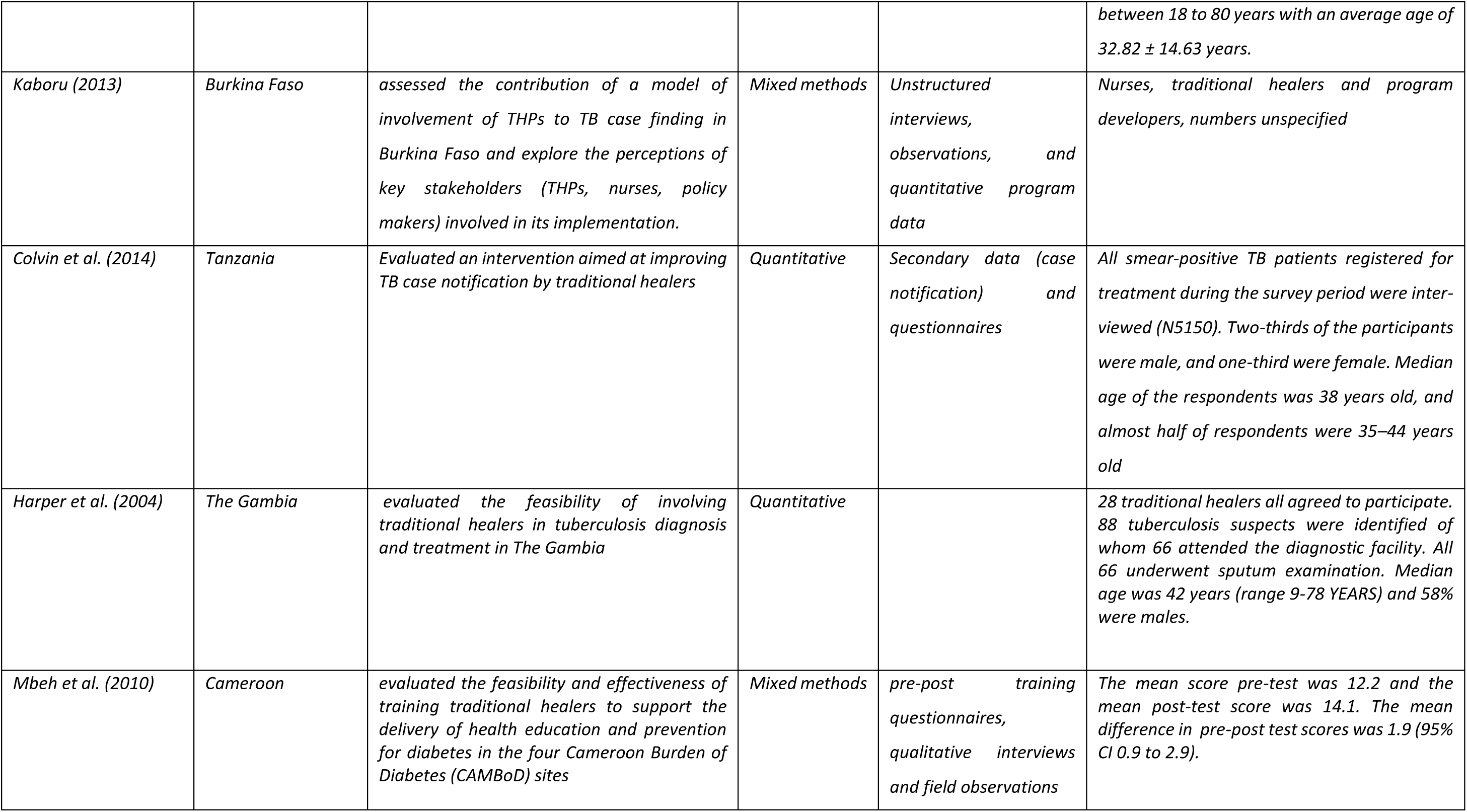
Description of Articles on Collaborative Interventions and Programs.

All five studies focusing on referral found some positive outcomes. In all studies there was an increase in the number of referrals by traditional healers^26–28,50,51^. Colvin et al. noted variation in referral rates over the two year monitoring, however, it was unclear if these were due to variations in presentation of tuberculosis or usage of referral tools by traditional healers^26^. Harper noted that the majority of their traditional healers continued referring patients after 1 year, those who stopped cited travel distances as their barrier. ^51^

Three studies measured the effect of educational interventions on knowledge and in all three traditional healers reported improved knowledge on diabetes prevention^30^, tuberculosis^51^ and HIV^52^. One study explored the participants perceptions on the integrated program to improve tuberculosis case finding. They found that the nurses involved were open to collaborating with traditional healers. For their part, traditional healers were surprised at the ease of the collaborative process and felt respected by the health workers. However, they felt they had the ability to perform more complex tasks^27^.

In Gyasi et al. 2017, the example of a system level attempt to integration, the authors explored perspectives on Ghana’s process of integrating traditional and biomedical practices. They included three stakeholder groups (biomedical professionals, traditional healers, and service users). They found that all groups felt the collaboration was needed but support was strongest among traditional healers and service users. Despite the support all participants were aware of gaps in term of training, registration, and regulation^29^.

### Models of Collaboration

In our introduction, we discussed the D’Amour et al. framework as an approach to classify types of collaboration: sharing, partnership, interdependency and power. We found discussions of all four constructs, but examples of only the first two levels of sharing and partnership.

### Sharing

We found that health care philosophy was substantially different between traditional healers and biomedical professionals. This philosophical divide varies by of traditional healer. Traditional healers who identified as spiritualists, were more likely to have very different philosophies, as compared to those who identified as bonesetters. Some biomedical professionals expressed more concern at collaboration with healers focused on the spiritual or religious aspects of healing. However, there were also biomedical professionals saw value in the spiritual healers as a support system for patients receiving biomedical treatment.

There was some consensus that both traditional healers and biomedical professionals shared the same values and wanted to improve patients’ health. However, Krah et al. discussed a particular problem with biomedical professionals being outsiders who rarely understood or shared the communities’ values. In Lampiao et al, all traditional healers believed they had the same goals as biomedical professionals. However, not all biomedical professionals believed they shared goals with traditional healers.

### Partnership

With regards to partnership, we found that the two most discussed attributes were ‘mutual trust and respect’ and ‘awareness and values’. The main finding was that trust was the most significant barrier to collaboration. This was often captured as traditional healers feeling that biomedical professionals did not trust or respect them for their knowledge, skills, and professionalism. This was exemplified by the one-way nature of referrals. However, there were also mentions of traditional healers not trusting biomedical professionals and their methods. In general, discussions on collaboration focused on traditional healers adopting the biomedical professionals’ ways of working. Biomedical professionals, provided support for these claims, often sharing their distrust of traditional healers’ methods and medications. Some reasons for the distrust are well-documented, including the lack of transparency in the development and ingredients of medications and healing practices.

Traditional healers demonstrated a greater acknowledgment of values of biomedicine, illustrated by instances of directing patients to health facilities for issues such as excessive bleeding and integrating biomedical medication into traditional healing practices. In contrast, biomedical professionals exhibited less certainty regarding the value of traditional healers and their methods, generally discouraging patients from seeking care from them. We also noticed some interesting contextual variations. For example, Upvall et al. study on nurses’ perceptions of collaboration revealed that those affiliated with mission and government hospitals in urban settings tended to be more dismissive of traditional healers than those in rural areas. Conversely, nurses in rural areas, particularly those associated with non-governmental organizations or private hospitals, displayed a more open attitude towards collaborating with traditional healers^36^.

### Interdependency and Power

There was no evidence of interdependence of general power sharing in any of the seven interventions studied.

## DISCUSSION

The aim of this review was to synthesise the literature on collaboration between traditional healer’s and biomedical professional including relevant interventions. Reflecting on included articles through the lens of D ’Amour et al. collaboration model, we found clear indicators related to sharing and partnership. However, no articles had elements of the constructs of interdependency and power.

Our review found that there are mostly positive attitudes towards the idea of collaboration between traditional healers and biomedical professionals. However, there is an evident gap between attitudes and system level collaboration. The seven articles which attempted to foster collaborative working agreements all reported positive outcomes. Yet none of these studies challenged existing power dynamics and hierarchies. They all involved traditional healers supporting biomedical professionals through screening or referrals. We would argue that true collaboration between the systems requires that both groups learn to value each other and gain an appreciation for the methods and epistemologies of the other. We think it would also involve cross-referral of patients across both actor groups. For example, a bio-medical practitioner may back refer a village patient to the traditional healer for ongoing maintenance of blood pressure control. If side effects occur or the blood pressure is inadequately controlled, then the traditional healer can refer the patient back to the specialist. The patient could then benefit from bio-medical care while obtaining ongoing psychological/spiritual benefit from regular contact with the traditional healers.

Comparing our findings with the existing literature on collaboration between traditional healers in mental health conditions^11,53–55^, we noticed a possible difference in trust across those catering to mental vs physical health. In studies that examined collaboration of health workers on mental health conditions, traditional healers were more likely to be dismissive of biomedical professionals when it came to their abilities to heal mental health ailments. This could be attributed to their different understandings of illness. Traditional healers tended to believe more strongly in the spiritual nature of mental illnesses. Whereas for more physical ailments included in our review, traditional healers seemed more open to utilising biomedical medications and processes.

Overall, we found that the majority of articles on traditional healers came from South Africa. South Africa has an established policy, indicating perhaps the importance of policy in promoting work on collaboration between traditional healers and biomedical professionals. However, there were no noticeable differences between attitudes in collaboration in South Africa and other countries with more recent policies. It is also important to note that a number of countries do have policies on integration, for example four of the fourteen Southern African countries (cited above). However, the policies have been criticised for weaknesses such an delays in enacting legislation and problems defining traditional practitioners^56^.

We also note that while some included articles addressed perceptions about general integration of traditional healers into primary care, intervention studies only addressed single health conditions. We think given the overall interest of traditional healers to work with biomedical professionals, the understanding of biomedical professionals concerns and the effectiveness of trained traditional healers, there is an opportunity to develop multi-condition models on collaboration between the systems.

## Data Availability

All data produced in the present work are contained in the manuscript

